# Evaluating Inadequate Therapy Response in Ulcerative Colitis Adult Patients: A Retrospective Analysis of German Health Claims Data on Advanced Therapy Initiation

**DOI:** 10.1101/2024.07.22.24310495

**Authors:** Axel Dignass, Irina Blumenstein, Carolina Schwedhelm, Katrin Strassen, Leonie Berger, Sophie Marquardt, Anna Seiffert, Nataliia Kulchytska, Ivonne Hänsel, Alexa Benson, Agnes Kisser

## Abstract

**Background:** The treatment landscape for active ulcerative colitis (UC) is rapidly evolving and current real-world evidence on response to advanced therapy is limited. This study aimed to determine indicators of inadequate therapeutic response among patients with UC in Germany initiating advanced therapy.

**Methods:** This retrospective analysis used German claims data (2015-2022) from adult patients (≥18 years). Prevalence and incidence of UC (ICD-10-GM: K51.X) were estimated. Inadequate response to therapy was evaluated in patients initiating advanced therapy based on eight predefined indicators observed for 12 months following index treatment prescription.

**Results:** Mean UC patient age in 2016-2022 ranged from 49.6 to 51.5 years, 47.6%-48.3% were female. Administrative prevalence ranged from 0.45% in 2016 to 0.53% in 2022. Number of patients initiating advanced treatment ranged from 157 to 347 across the study years (3.2%-4.9% of overall treated study population). On average from 2016-2021, 78.8% had inadequate response in the 12 months following index treatment. Common indicators included prolonged use of corticosteroids (46.2%) and augmentation with conventional therapies (43.9%).

**Conclusions:** Adult UC patients showed a high prevalence of inadequate response to advanced therapies. Our findings reveal a need for improved UC advanced therapy options, providing insight into inadequate response patterns. This may help identify patients who could benefit from a change in therapy to improve long-term outcomes.

## Introduction

Ulcerative colitis (UC) is a chronic inflammatory bowel disease affecting the colon and rectum, causing significant morbidity and impaired quality of life^1,2^. In Germany, approximately 150,000-160,000 individuals live with UC^3,4^, with a standardized prevalence of 5.3 per 1,000 in 2019^5^. Despite increased availability of advanced therapies, including biologics and JAK inhibitors, treatment outcomes for UC have not significantly improved. Many patients still experience inadequate therapeutic response, requiring frequent dose escalations and therapy switches^6^. This affects individual outcomes and increases healthcare costs^7^. A recent study highlighted that patients exposed to multiple prior therapies are more likely to respond inadequately to subsequent treatments^7^. Given the substantial burdens of active UC, better treatment options are critically needed to achieve and maintain disease remission without longterm corticosteroid (CS) use^2^.

This study aimed to complement previous studies of adult prevalent and incident UC cases using real-world German claims data and identify cases of inadequate response in patients initiating advanced treatment.

## Methods

This non-interventional, longitudinal, retrospective cohort study utilized anonymized German claims data from the German Analysis Database for Evaluation and Health Services Research (DADB) – managed by Gesundheitsforen Leipzig GmbH – to examine moderate to severe UC patients who had an inadequate response to initial advanced treatment. The DADB includes data from 15 statutory health insurers (SHI) from 2013-2022; representing ^~^4.1 million individuals and is representative of the broader German population.

The study period spanned January 1, 2015, to December 31, 2022. Inclusion criteria encompassed individuals ≥12 years with at least two secondary diagnoses in two quarters within one calendar year or one inpatient primary diagnosis of UC (ICD-10-GM: K51.X). Due to low adolescent patient numbers, this study focused on adults (≥ 18 years). Prevalent and incident patients required observable data for at least one year and two years, respectively, before their UC diagnosis and subsequent therapy initiation. Exclusion criteria included Crohn’s disease, indeterminate colitis, or other conditions warranting advanced therapy. Patients were categorized by their initiation of advanced UC therapy (adalimumab, golimumab, infliximab, vedolizumab, ustekinumab, tofacitinib, upadacitinib, filgotinib, ozanimod). Patients without prescriptions for advanced UC therapy in the 12 months prior were considered as ‘treatment-naive’ (i.e., starting first advanced treatment); those prescribed a different advanced therapy agent were considered as ‘treatment-experienced’ (i.e., changing advanced treatment). Inadequate response to index treatment was evaluated among patients initiating first advanced treatment based on indicators observed for 12 months following the initial prescription.

We evaluated indicators described by Bokemeyer et al.^7^, in addition to prescriptions for anti-diarrheal medications (Table 1). Patient numbers, proportions for prevalence, incidence, clinical characteristics, and inadequate therapy response (assumed when patients exhibited ≥1 indicator) were estimated per calendar year. A sensitivity analysis was performed for multifactorial inadequate response (i.e., ≥2 and ≥3 indicators). Due to the 12-month observation period, inadequate response analysis is not available for 2022 index therapies. The study analysis was carried out in Microsoft SQL Server and in R.

**TABLE 1:** Indicators of inadequate response to UC therapy.

| Indicator | Definition |
| --- | --- |
| Dose escalation | Average prescribed daily dose of index agent was $\geq 150\%$ of the standard maintenance dose over at least 3 prescriptions |
| Therapy switch | Switch from the index agent to another advanced therapy and no refill of the index agent within 180 days |
| Augmentation | Prescription of a conventional therapy (aminosalicylates, immunosuppressives, or CS) in addition to the index therapy |
| Prolonged use of CS | At least two outpatient CS prescriptions |
| Discontinuation | A gap of $> 60$ days between consuming the previous index prescription's supply and the next index prescription |
| UC-related surgery | At least one inpatient or outpatient procedure (intestinal incision, excision, resection and anastomosis; other intestinal procedures; rectal procedures; anus procedures) |
| UC-related hospitalization | At least one primary UC or UC-related diagnosis (abscesses, acute and subacute iridocyclitis, cholangitis, erythema nodosum, pyoderma gangraenosum, primary sclerosing cholangitis) |
| Anti-diarrheal medication Rx | At least one prescription (intestinal adsorbents, electrolytes with carbohydrates, motility inhibitors, or microbial anti-diarrheals) |

CS, corticosteroids; RX, prescription; UC, ulcerative colitis.

## Results

### Patient selection and characteristics

Identified UC patients with full insurance coverage the calendar year prior to UC diagnosis ranged from 9,295 in 2016 to 12,613 in 2022, corresponding to 271,752 and 330,768 in the German SHI population. In 2021, the treated UC patient population was 7,077, with 6,859 patients having an identifiable index treatment agent (Table 2). The overall treated study population in 2021 averaged 51.0 years (± 16.3), while patients initiating or changing advanced treatment were younger (42.7 and 44.7 years, respectively) (Table 3). Gender distribution was balanced, with females constituting 47.6%-48.3% of the overall treated population. In 2021, 347 patients initiated first advanced treatment and 54 changed advanced treatment (Table 3), corresponding to 8,499 and 1,278 German SHI patients, respectively. The predominant index treatment was TNF-α-inhibitors, followed by integrin-inhibitors (2016-2021 range: 86.3%-53.3% and 13.7%-38.0%, respectively) (Table 4).

**TABLE 2:** Patient selection process.

| Selection step | 2016 | 2017 | 2018 | 2019 | 2020 | 2021 | 2022 |
| --- | --- | --- | --- | --- | --- | --- | --- |
| 1) Persons in the DADB database | 2,216,982 | 2,364,446 | 2,414,261 | 2,441,254 | 2,424,861 | 2,426,311 | 2,458,016 |
| ▼ |  |  |  |  |  |  |  |
| 2) Diagnosis M2Q K51 | 9,833 | 10,914 | 11,560 | 12,086 | 12,180 | 12,767 | 13,015 |
| ▼ |  |  |  |  |  |  |  |
| 3) Full health insurance coverage calendar year prior to year of UC diagnosis | 9,295 | 9,981 | 11,072 | 11,686 | 11,871 | 12,406 | 12,613 |
| ▼ |  |  |  |  |  |  |  |
| 4) At least one prescription of UC therapy after UC index diagnosis | 5,830 | 6,589 | 7,552 | 8,153 | 8,459 | 8,799 | 9,163 |
| ▼ |  |  |  |  |  |  |  |
| 5) No diagnosis of Crohn's disease and other specific diseases within 365 days preceding index treatment date | 4,976 | 5,504 | 6,196 | 6,603 | 6,818 | 7,077 | 7,277 |
| ▼ |  |  |  |  |  |  |  |
| **6) Overall treated study population** | 4,976 | 5,504 | 6,196 | 6,603 | 6,818 | 7,077 | 7,277 |
| ▼ |  |  |  |  |  |  |  |
| 7) Pre-observation period of at least 12 months before index treatment date | 4,976 | 5,504 | 6,157 | 6,558 | 6,776 | 7,044 | 7,236 |
| ▼ |  |  |  |  |  |  |  |
| 8) Index treatment agent can be determined (multiple prescriptions on index date excluded) | 4,726 | 5,256 | 5,919 | 6,310 | 6,552 | 6,859 | 7,034 |
| ▼ |  |  |  |  |  |  |  |
| **9a) patients with first advanced treatment** | 157 | 222 | 257 | 305 | 295 | 347 | 335 |
| **9b) patients with change of advanced treatment** | 25 | 25 | 18 | 53 | 49 | 54 | 49 |

DADB, database for evaluation and health services research; M2Q K51, a confirmed outpatient or inpatient secondary UC diagnosis in at least two quarters or as one inpatient primary diagnosis; UC, ulcerative colitis.

**Table 3:**
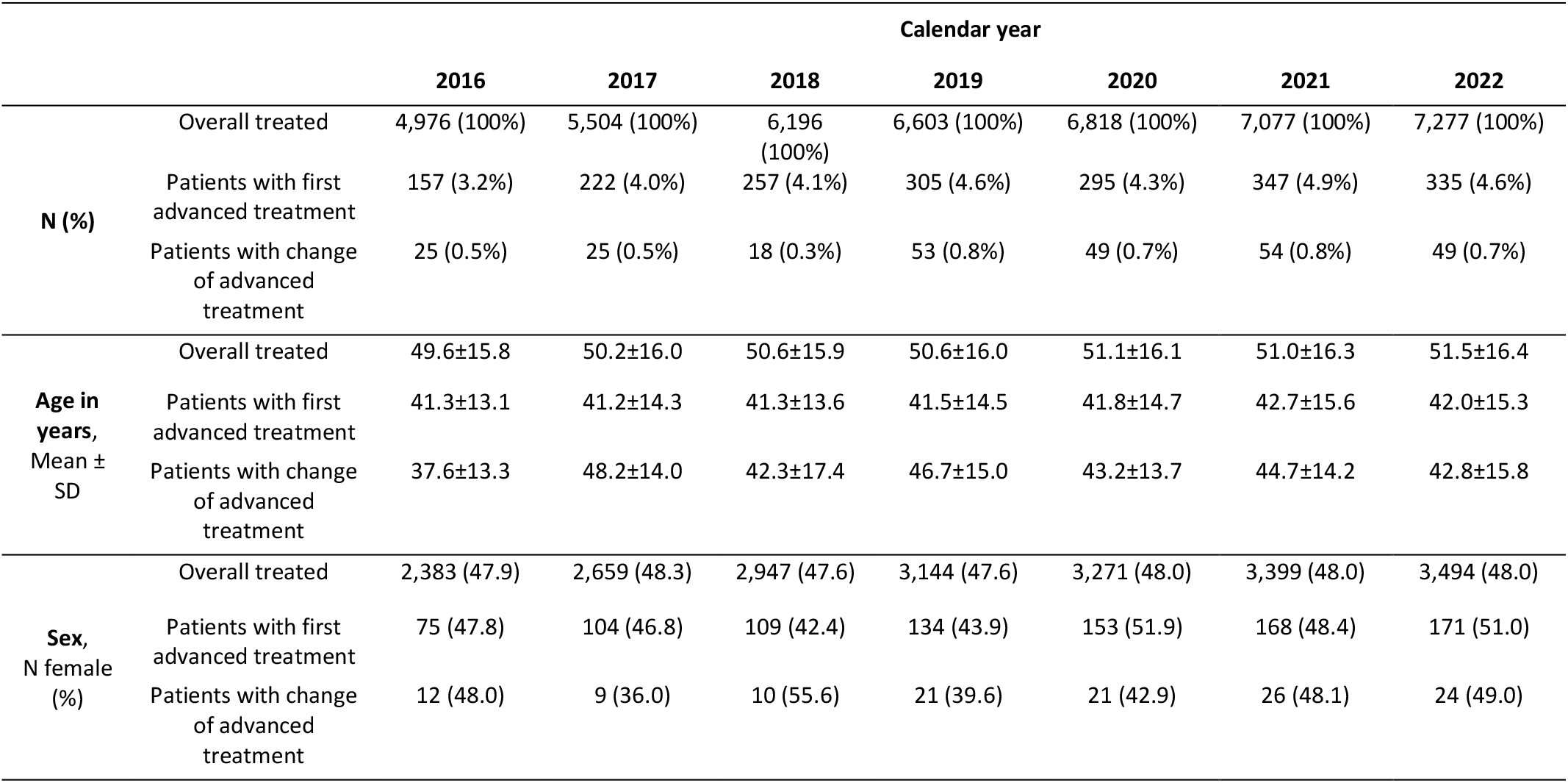
Patient characteristics of the overall treated patient population.

|  |  | Calendar year |  |  |  |  |  |  |
| --- | --- | --- | --- | --- | --- | --- | --- | --- |
|  |  | 2016 | 2017 | 2018 | 2019 | 2020 | 2021 | 2022 |
| **N (%)** | Overall treated | 4,976 (100%) | 5,504 (100%) | 6,196 (100%) | 6,603 (100%) | 6,818 (100%) | 7,077 (100%) | 7,277 (100%) |
|  | Patients with first advanced treatment | 157 (3.2%) | 222 (4.0%) | 257 (4.1%) | 305 (4.6%) | 295 (4.3%) | 347 (4.9%) | 335 (4.6%) |
|  | Patients with change of advanced treatment | 25 (0.5%) | 25 (0.5%) | 18 (0.3%) | 53 (0.8%) | 49 (0.7%) | 54 (0.8%) | 49 (0.7%) |
| **Age in years, Mean ± SD** | Overall treated | 49.6±15.8 | 50.2±16.0 | 50.6±15.9 | 50.6±16.0 | 51.1±16.1 | 51.0±16.3 | 51.5±16.4 |
| | Patients with first advanced treatment | 41.3±13.1 | 41.2±14.3 | 41.3±13.6 | 41.5±14.5 | 41.8±14.7 | 42.7±15.6 | 42.0±15.3 |
| | Patients with change of advanced treatment | 37.6±13.3 | 48.2±14.0 | 42.3±17.4 | 46.7±15.0 | 43.2±13.7 | 44.7±14.2 | 42.8±15.8 |
| **Sex, N female (%)** | Overall treated | 2,383 (47.9) | 2,659 (48.3) | 2,947 (47.6) | 3,144 (47.6) | 3,271 (48.0) | 3,399 (48.0) | 3,494 (48.0) |
|  | Patients with first advanced treatment | 75 (47.8) | 104 (46.8) | 109 (42.4) | 134 (43.9) | 153 (51.9) | 168 (48.4) | 171 (51.0) |
|  | Patients with change of advanced treatment | 12 (48.0) | 9 (36.0) | 10 (55.6) | 21 (39.6) | 21 (42.9) | 26 (48.1) | 24 (49.0) |

**Table 4:** Number of patients with first advanced treatment by substance class of index agent.

|  | N (%) per calendar year |  |  |  |  |  |
| --- | --- | --- | --- | --- | --- | --- |
|  | 2016 | 2017 | 2018 | 2019 | 2020 | 2021 |
| Total<sup>a</sup> | 153 (100.0%) | 206 (100.0%) | 251 (100.0%) | 288 (100.0%) | 276 (100.0%) | 334 (100.0%) |
| **Substance class of index agent** |  |  |  |  |  |  |
| TNF-α-inhibitor (Adalimumab, Infliximab, Golimumab) | 132 (86.3%) | 158 (76.7%) | 190 (75.7%) | 193 (67.0%) | 163 (59.1%) | 178 (53.3%) |
| Integrin-inhibitor (Vedolizumab) | 21 (13.7%) | 46 (22.3%) | 56 (22.3%) | 73 (25.3%) | 87 (31.5%) | 127 (38.0%) |
| IL12/IL23-inhibitor (Ustekinumab) | N<5<sup>b</sup> | N<5 | N<5 | 8 (2.8%) | 20 (7.2%) | 20 (6.0%) |
| JAK-inhibitor (Tofacitinib, Upadacitinib, Filgotinib) | N<5 | N<5 | N<5 | 14 (4.9%) | 6 (2.2%) | 9 (2.7%) |
| Sphingosine-1-phosphate receptor modulator (Ozanimod) | N<5 | N<5 | N<5 | N<5 | N<5 | N<5 |

<sup>a</sup> Number of patients with follow-up period of at least 12 months after prescription of index therapy. Patients excluded due to incomplete follow-up time (< 12 months): N=4 (2016), 16 (2017), 6 (2018), 17 (2019), 19 (2020), 13 (2021).
<sup>b</sup> Patient number under N=5 not available due to data protection regulations *Adult UC prevalence and incidence*

Total UC prevalence increased slightly from 0.45% (95%CI, 0.44% to 0.46%) to 0.53% (95%CI, 0.52% to 0.54%), and prevalence of UC-treated patients from 0.31% (95%CI, 0.30% to 0.32%) to 0.39% (95%CI, 0.38% to 0.39%) in 2016 and 2022, respectively (2016-2022 average: 0.49% and 0.35%, respectively). Total and UC-treated patient incidence remained around 0.04% and 0.025%, respectively. Incidence estimates were slightly lower in 2022 (total UC incidence: 0.034%, 95%CI: 0.032-0.037%) compared to 2017 and 2018 (Figure 1A). Adult UC prevalence in 2021 increased with age, peaking in the 60-64 age group (Figure 1B). Gender differences revealed higher prevalence among younger females, whereas males exhibited higher prevalence in middle and older age groups (Figure 1B).

**FIGURE 1:**
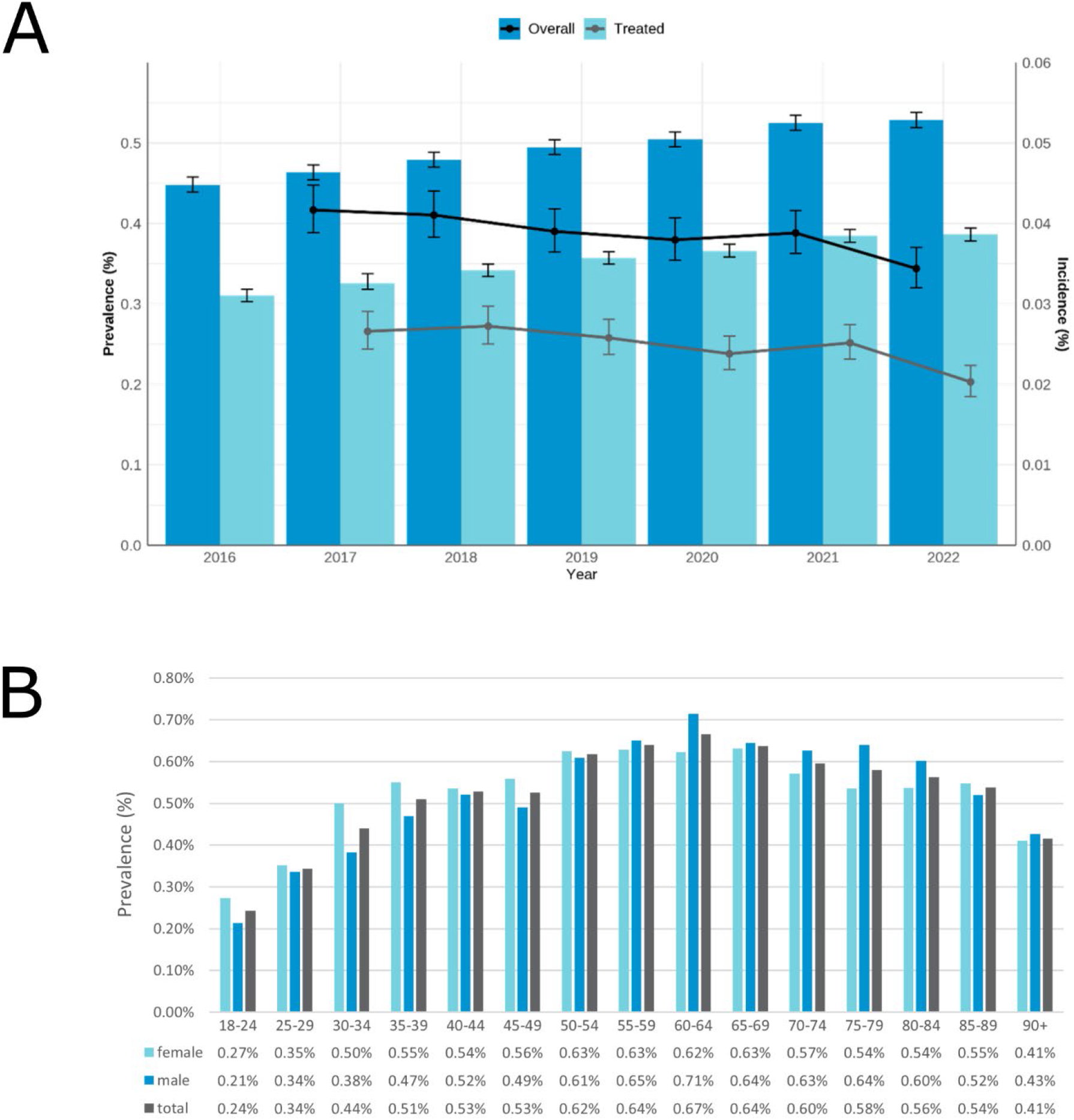
Adult prevalence and incidence of UC (overall UC population and UC-treated population) in the German SHI population by year. Prevalence is indicated by the bar plots and incidence is indicated by the line plots [A], and UC prevalence by age and gender group in the German SHI population, 2021 [B].

### Inadequate response to first advanced treatment

Among patients initiating their first advanced treatment in 2021, 71.2% exhibited at least one indicator for inadequate response (2016-2021 average 78.8%), while 48.9% (2016-2021 average 55.3%) exhibited a multifactorial inadequate response of ≥2 indicators, and 25.7% (2016-2021 average 31.4%) with ≥3 indicators (Figure 2A). On average for index treatments from 2016-2021, the most common sign of inadequate response was prolonged use of CS (46.2%), followed by augmentation with conventional therapies (43.9%) (Figure 2B). Less common indications included dose escalation (8.8%), UC-related surgery (7.2%), and anti-diarrheal prescriptions (6.4%). Overall inadequate response rates and individual indicators were generally lower for years 2020 and 2021 (Figure 2).

**FIGURE 2:**
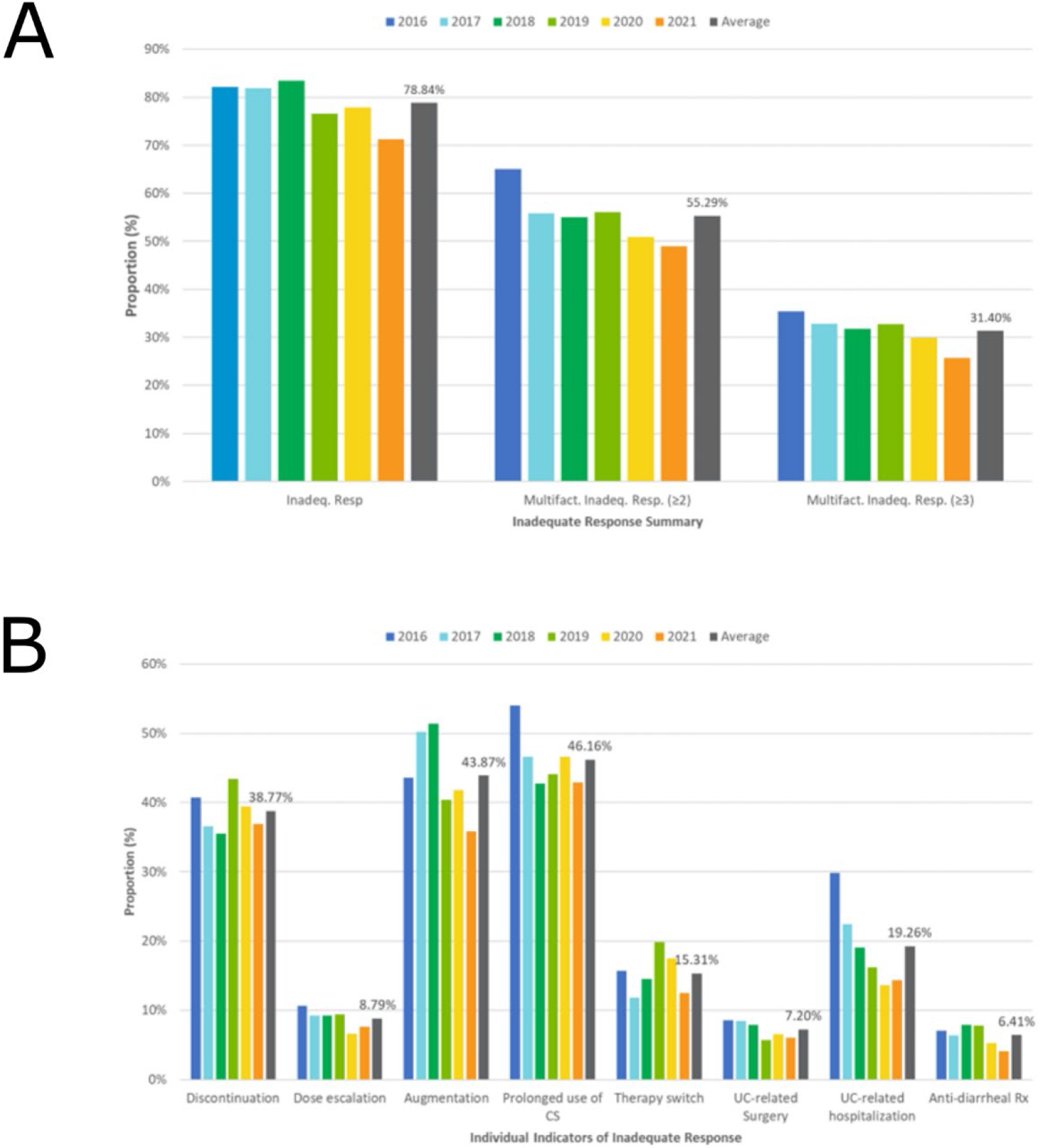
Proportion of patients showing inadequate response to first advanced treatment based on presence of ≥1, ≥2, or ≥3 indicators (multifactorial inadequate response) [A], and proportion of patients showing individual indicators of inadequate response [B]. Patients excluded due to incomplete follow-up time (< 12 months): N=4 (2016), 16 (2017), 6 (2018), 17 (2019), 19 (2020), 13 (2021). CS, corticosteroids; Rx, prescription; UC, ulcerative colitis.

## Discussion

This study provides valuable real-world evidence on the adult prevalence and incidence of UC in Germany and on response patterns to the first advanced therapy. Our findings reveal a high prevalence of inadequate response to the first advanced therapy among adult patients, highlighting the underlying patterns of inadequate UC therapy response in the current treatment landscape. This evidence may help the scientific and medical community identify patients for whom a change in therapy might be particularly beneficial, especially those exhibiting multifactorial inadequate response.

Our findings of increasing prevalence over the years are in line with previous studies^7-9^, and the total UC prevalence of 0.45%-0.53% was consistent with the 2019 estimation of Mueller (0.53%)^5^. The decreasing trend in UC incidence after the onset of the pandemic could be explained by fewer doctors and hospital visits^10,11^. Previous studies reflect a different treatment landscape, as new treatments have become available in the last decade. Nevertheless, our findings are consistent with a previous German claims study by Bokemeyer et al.^7^, and a US claims study by Rubin et al.^9^, reporting a suboptimal treatment response of 75% and 81%, respectively, after 12 months follow-up. The most common indicators of inadequate response were prolonged CS use (46.2%) and augmentation with conventional therapies (43.9%), slightly higher than found in Bokemeyer’s report^7^ (35.8% and 30.5%, respectively), while augmentation was consistent with Rubin’s findings^9^. We observed that 38.8% of patients experienced therapy discontinuation. Reasons for discontinuation could be suboptimal therapy or safety, however, such information is not available. These findings are consistent with Bokemeyer’s report of a 37.7% therapy discontinuation rate at one year^7^, and Mevius’s finding that 30% to 40% of patients discontinued biologics within one year^8^. Overall, these results underscore the challenges in managing UC, despite the availability of advanced treatment options.

Our study’s strength lies in its utilization of a comprehensive, anonymized, and representative claims database. Another strength is the study’s adaption to the current treatment landscape, including all advanced therapies approved by the European Medicines Agency (EMA) for UC in Germany. However, the timeframe must be understood in the context of the evolving UC advanced treatment landscape and the COVID-19 pandemic. The study’s retrospective design and reliance on claims data may not capture all clinical nuances, limiting data interpretability. Individual follow-up is not possible, and miscoding cannot be excluded.

In conclusion, this study highlights the urgent need for new advanced therapies in Germany to address the high inadequate response rate in UC. Despite multiple advanced therapeutic options, patients do not achieve adequate disease control, necessitating ongoing research and development of more effective treatments.

## Data Availability

The datasets generated and/or analyzed during the current study are not publicly available duet to restriction of data provider. Aggregated data were provided by the Gesundheitsforen Leipzig GmbH.

## Author contributions

AD, IB, CS, LB, SM, NK, IH and AK were involved in the design of the study. KS and AS were involved in the data analysis. AD, IB, CS, KS, LB, NK, IH, AB, and AK were involved in the data interpretation. AB wrote the manuscript, and AD, IB, CS, LB, AS, NK, IH, and AK provided critical revisions to the manuscript. All authors read and approved the final abstract.

